# Trend Analysis and Forecasting of COVID-19 outbreak in India

**DOI:** 10.1101/2020.03.26.20044511

**Authors:** Rajan Gupta, Saibal K Pal

**Affiliations:** Deen Dayal Upadhyaya College, University of Delhi, Delhi, India; DRDO, Delhi, India

**Keywords:** COVID-19, India, Novel Coronavirus, nCov, Trend Analysis, Forecasting, Disease Outbreak, ARIMA, Holt’s Trend, Exponential Smoothing, Predictive Analytics

## Abstract

COVID-19 is spreading really fast around the world. The current study describes the situation of the outbreak of this disease in India and predicts the number of cases expected to rise in India. The study also discusses the regional analysis of Indian states and presents the preparedness level of India in combating this outbreak. The study uses exploratory data analysis to report the current situation and uses time-series forecasting methods to predict the future trends. The data has been considered from the repository of John Hopkins University and covers up the time period from 30^th^ January 2020 when the first case occurred in India till the end of 24^th^ March 2020 when the Prime Minister of India declared a complete lockdown in the country for 21 days starting 25^th^ March 2020. The major findings show that number of infected cases in India is rising quickly with the average infected cases per day rising from 10 to 73 from the first case to the 300^th^ case. The current mortality rate for India stands around 1.9. Kerala and Maharashtra are the top two infected states in India with more than 100 infected cases reported in each state, respectively. A total of 25 states have reported at least one infected case, however only 8 of them have reported deaths due to COVID-19. The ARIMA model prediction shows that the infected cases in India may reach up to 700 thousands in next 30 days in worst case scenario while most optimistic scenario may restrict the numbers up to 1000-1200. Also, the average forecast by ARIMA model in next 30 days is around 7000 patients from the current numbers of 536. Based on the forecasting model by Holt’s linear trends, an expected 3 million people may get infected if control measures are not taken in the near future. This study will be useful for the key stakeholders like Government Officials and Medical Practitioners in assessing the trends for India and preparing a combat plan with stringent measures. Also, this study will be helpful for data scientists, statisticians, mathematicians and analytics professionals in predicting outbreak numbers with better accuracy.

## I. Introduction

COVID-19 [1][2] is spreading really fast around the world. COVID-19 is a respiratory infection with common signs that include respiratory symptoms, fever, cough, shortness of breath, and breathing difficulties. In more severe cases, infection can cause pneumonia, severe acute respiratory syndrome, kidney failure, and death. It has infected more than 400 thousand people all around the world till the end of the day on 24^th^ March 2020. The risk of severe complications from COVID-19 is higher for certain vulnerable populations, particularly people who are elderly, frail, or have multiple chronic conditions. Although the situation is worse in European and American region after it got stabilized in China, the conditions in South Asia are deteriorating fast.

India is the leading region in South Asian region which is the second most populous country in the world after China. Uncontrolled pandemic in India can lead to the impact of almost 1/6^th^ population of the world. India holds a significant value on the world map with respect to trade, economy, defense, culture, entertainment, outsourcing workforce, manufacturing and services. Indians are present in almost all the regions of the world and are working towards strengthening the work processes and economies of the world. It is imperative that the trends and forecasts for Indian region must be studied well so that effective strategies can be drawn for the region. Since the mechanisms of COVID-19 spreading are not completely understood, the number of infected people is large and the effects of containment are evaluated essentially on an empirical basis. Therefore, a more quantitative analysis of the epidemic spreading can be interesting.

There have been many modelling approaches presented by various mathematicians and researchers in recent time for different countries. Models for China [3-7], Italy [8-11], France [8][12], USA [13-15], and South Korea [16-17]. For Indian region, the studies are very less till now and some notable work has been done by people from ICMR [18] and other researchers [19-21]. Except from the mathematical modelling presented by the practitioners from Indian Council of Medical Research, there are no prediction and forecasting models available in the Indian context.

From analysis point of view, a lot of studies have been conducted using time series forecasting models like ARIMA [22-24] and Exponential smoothing [25-27]. These are standard techniques which gives a decent predictions and forecasts on time series data in quick time. These techniques have been selected based on their wide acceptance in the research community and quick implementation for the various stakeholders to act.

The objective of the current study is to analyze the COVID-19 outbreak situation in India and assess the trends in near future. Also the study aims to take an overview of the preparedness levels of this outbreak from Indian Government. The scope of this study is limited to building forecasting models for Indian region and uses time series based forecasting methods which are easy to build and easy to understand in these kind of critical conditions. The study does not include forecasting for any other nation suffering from COVID-19 outbreak.

## II. Techniques

The current analysis has been divided into various phases. Firstly, the trend analysis of number of infected cases and death cases has been assessed for India. Next the state wise analysis has been conducted for India showcasing various penetration levels and mortality rates for different states and union territories in the country. Thirdly, the comparative analysis of India with other SAARC nations have been done to assess the level of COVID-19 spread in south Asian region. Then, the two forecasting methods have been applied on the natural log transformed data collected from the John Hopkins University website [28]. The data has been considered for Indian region from 30-Jan-2020 onwards when the first case of COVID-19 was reported, up to 24^th^ March 2020, when complete lockdown of the nation has been imposed by the Government of India. Lastly, the medical infrastructure and preparedness level of India has been analyzed through various secondary reports and news articles in order to understand the seriousness of the forecasting numbers.

Based on the popular methods available for modeling and forecasting the time series data, ARIMA modeling and Exponential Smoothing methods have been implemented in this study. ARIMA (Auto-Regressive Integrated Moving Average) models [29] are, in theory, the most general class of models for forecasting a time series which can be made to be “stationary” by differencing (if necessary), perhaps in conjunction with nonlinear transformations such as logging or deflating (if necessary). A random variable that is a time series is stationary if its statistical properties are all constant over time. A stationary series has no trend, its variations around its mean have constant amplitude, and it wiggles in a consistent fashion, i.e., its short-term random time patterns always look the same in a statistical sense. A non-seasonal ARIMA model is classified as an ARIMA(p, d, q) model, where *p* is the number of autoregressive terms, *d* is the number of non-seasonal differences needed for stationarity, and *q* is the number of lagged forecast errors in the prediction equation. The forecasting equation is constructed as shown in eq. 1.

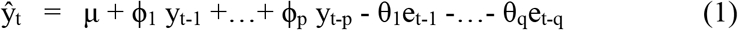

Here the moving average parameters (θ’s) are defined so that their signs are negative in the equation, following the convention introduced by Box and Jenkins. For the current study, we have considered p=1, d=1 and q=2 to construct the ARIMA (1,1,2) model using equation 2, as these for these values the model’s coefficients were computed to be most significant.

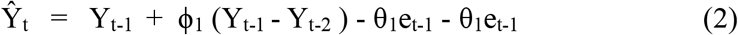

Since the time series forecasting using ARIMA should be done on a stationary series, the Dickey-Fuller test of stationarity was done to check the series. Rolling Mean and Rolling Standard Deviations were also calculated and the differences were calculated from the original data. Also the original data was log transformed to reduce the seasonality and trend in the original data series. Both ARIMA and LES were applied on the log transformed data, however the descriptive analysis was done on the original dataset.

Holt’s Linear Exponential Smoothing (LES) uses two smoothing constants to smoothen the level and trend in the data [30][31]. It is an important method if the data shows trend and not seasonality. In the current case, there is a clear upward trend in the number of infected cases from COVID-19 in India but seasonality is yet not a concern as the data is limited to a few days only. So this technique should work well for the current series. In the Holt’s LES, if the estimated level and trend at time *t-1* are *L*_*t-1*_ and *T*_*t-1*_, respectively, then the forecast for *Y*_*t*_ that would have been made at time *t-1* is equal to *L*_*t-1*_ + *T*_*t-1*_. When the actual value is observed, the updated estimate of the level is computed recursively by interpolating between *Y*_*t*_ and its forecast, *L*_*t-1*_ + *T*_*t-1*_, using weights of *α* and *1-α* as shown in equation 3.

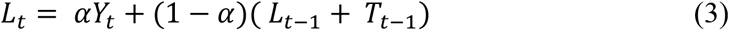

The change in the estimated level, namely *L*_*t*_ *-L*_*t-1*_, can be interpreted as a noisy measurement of the trend at time *t*. The updated estimate of the trend is then computed recursively by interpolating between *L*_*t*_ *-L*_*t-1*_ and the previous estimate of the trend, *T*_*t-1*_, using weights of *β* and *1-β* as shown in equation 4.

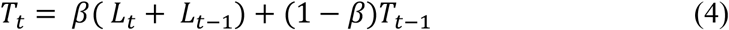

Finally, the forecasts for the near future that are made from time *t* are obtained by extrapolation of the updated level and trend as shown in equation 5.

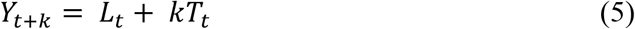

The interpretation of the trend-smoothing constant β is analogous to that of the level – smoothing constant α. Models with small values of β assume that the trend changes only very slowly over time, while models with larger β assume that it is changing more rapidly, where 0≤ α ≤1 is the level smoothing parameter, and 0≤ β ≤1 is the trend smoothing parameter. For long-term forecast, forecasting with Holt’s method will increase or decrease indefinitely into the future. In this case, we use the Damped trend method which has a damping parameter 0< ϕ <1 to prevent the forecast “go wild”. We implemented three variants of Holt’s method. In fit 1, we explicitly provide the model with the smoothing parameter α=0.8, β=0.2. In fit 2, we used an exponential model rather than a Holt’s additive model (which is default). In fit 3, we used a damped version of the Holt’s additive model but allowed the dampening parameter ϕ to be optimized while fixing the values for α=0.8, β=0.2. The values of α & β were set as per the trends seen in the original dataset. These values were obtained after iteratively developing the model.

All the codes have been implemented using Python 3.7 version in Jupyter Notebook. The data files have been downloaded from GitHub account of John Hopkins University in csv format.

## III. Analysis

### A. Trends in India

Like other countries, India is currently witnessing a steep rise in the number of cases on daily basis as shown in Figure 1.

**Figure 1.**
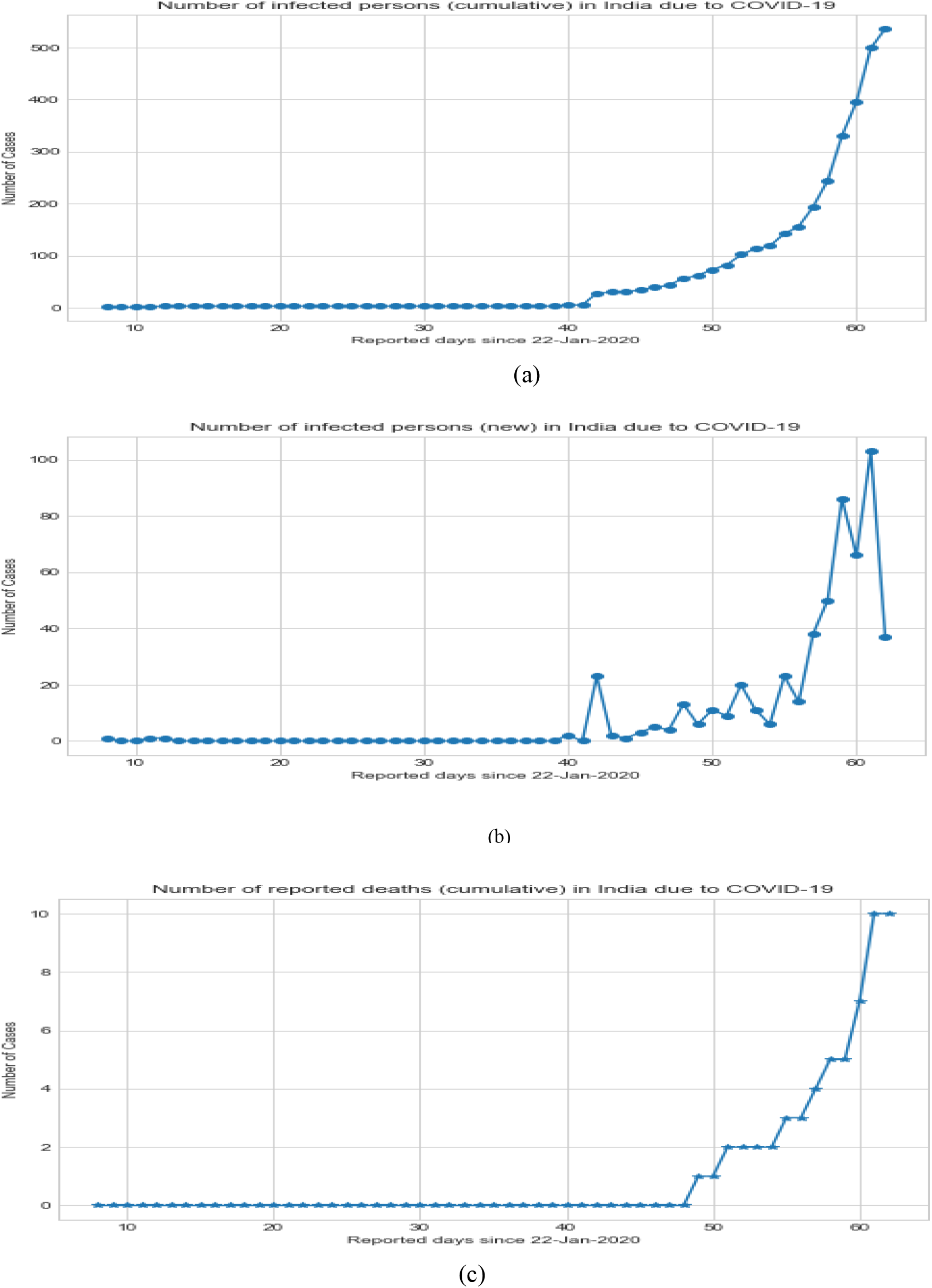
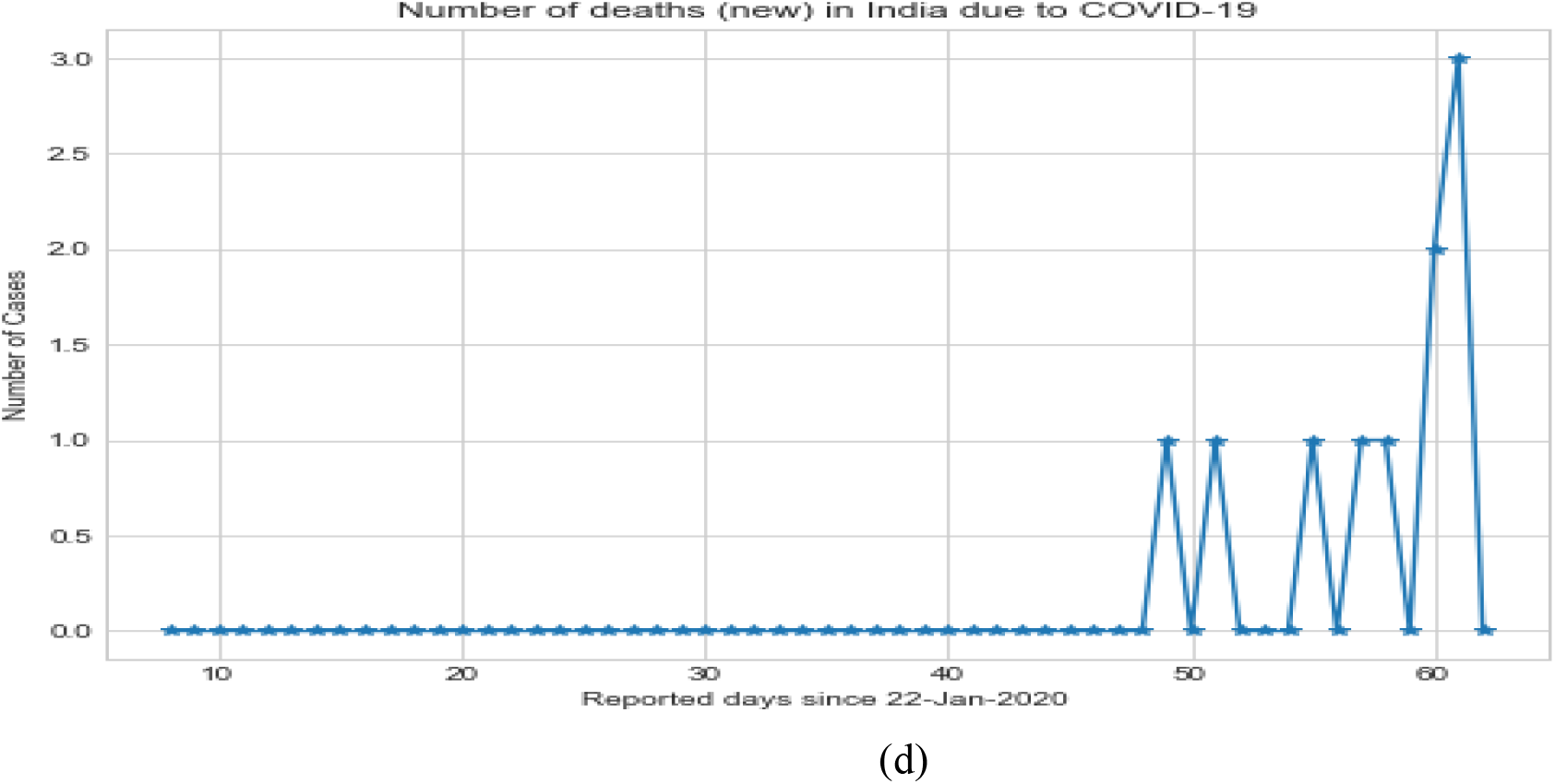
Trends for cases due to COVID-19 in India. (a) Cumulative number of infected cases. (b) New infected cases (c) Cumulative number of death cases (d) New death cases.

As seen from the figure and trends depicted by the dataset for COVID-19 outbreak, the cumulative number of infectious people as on 24^th^ March 2020 was 536 and there were 10 deaths reported for these infected people making it a mortality rate of almost 1.9%. This rate is much lower than the mortality rate at world level which is hovering around 4% and is very high in European region touching around 7-8%. The number of infected people increased in last one week as on 17^th^ March 2020 there were only 142 reported cases of infections and only 2-3 deaths were reported. Suddenly in one week, the number of infectious people and number of deaths increased by almost 3 times. Before entering the 3^rd^ week of March, the new cases of infections were almost constant and there were no major deaths. But by the end of second week of March, the trend line went upward very sharply and in the next 10 days majority of the damage has been done.

Figure 2 shows the trends and analysis of the number of cases reported on daily basis in India. Going by the trend, it can be seen that since registering the first case of COVID-19, it took 41 days in India to reach 50 infected patients and the average case per day is 9.75 since the reporting of first case. This is a normal trend but then it took a steep movement upwards where India went from 51 to 100 cases in next 5 days, followed by 1011-150 patients in another 5 days. The number of days went down to 3, 2 and 1 for the next 50 patients each from 151 to 300. After that it’s a matter of 1 or 2 days to regularly report 50 patients infected from COVID-19. There have been some milestones on finding the average case per day. Since reporting the first case till 24^th^ March 2020, the average case per day comes out to be 9.75 as in the initial days there were hardly any patients diagnosed with the virus. But this average kept on increasing with every case. Since reporting the 20^th^ case, average went to 25.29 cases per day, since 50^th^ case it was 32.87, since 100^th^ case it was 41.27, since 150^th^ case it was 41.27, since 200^th^ case it was 68.4, and since 250^th^ and 300^th^ case it was 73, respectively. There was a slight downfall after that as a complete one day lockdown was announced by the Government of India. This average came down to 70 which is almost constant for last 100 cases.

**Figure 2.**
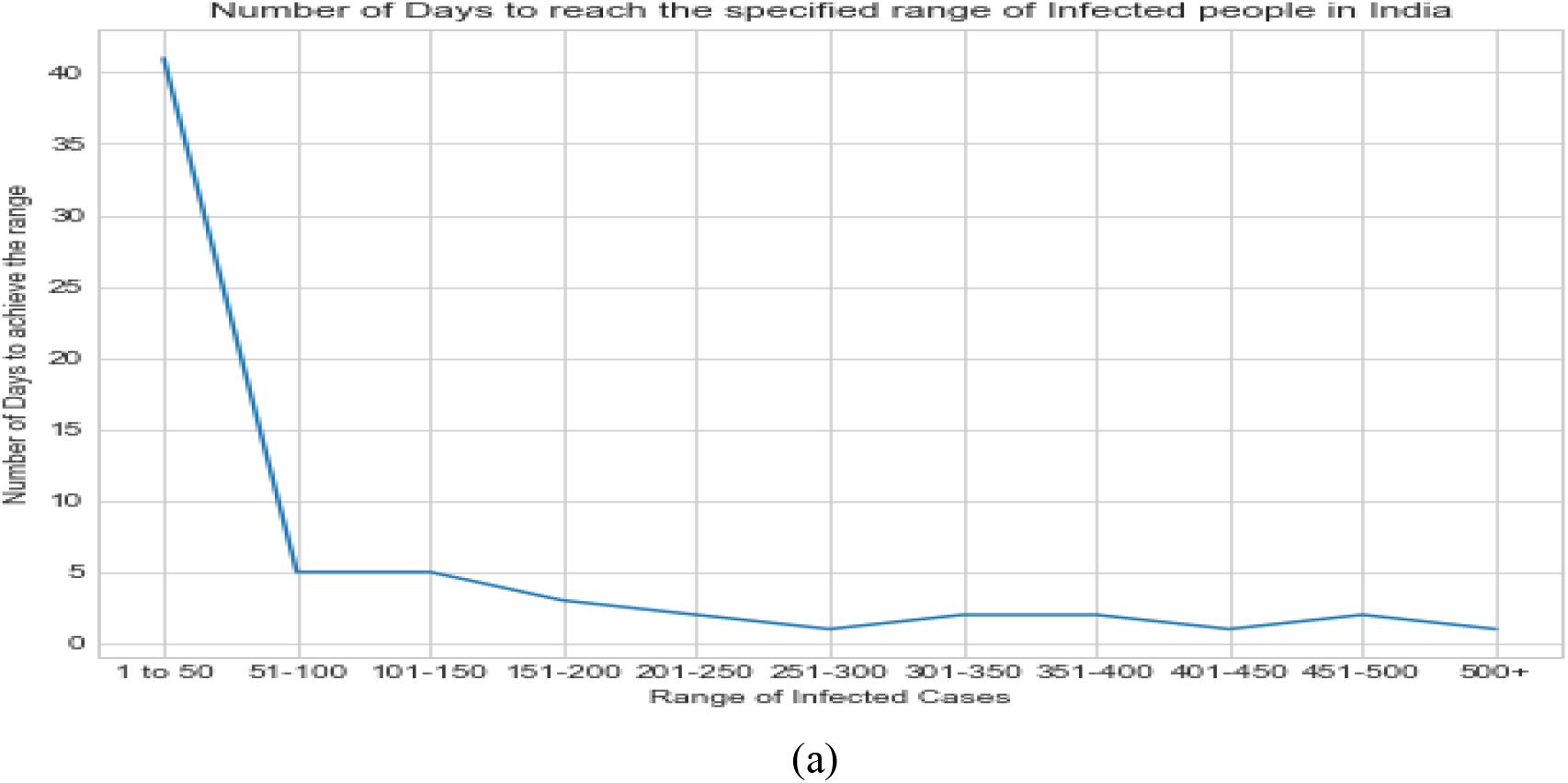
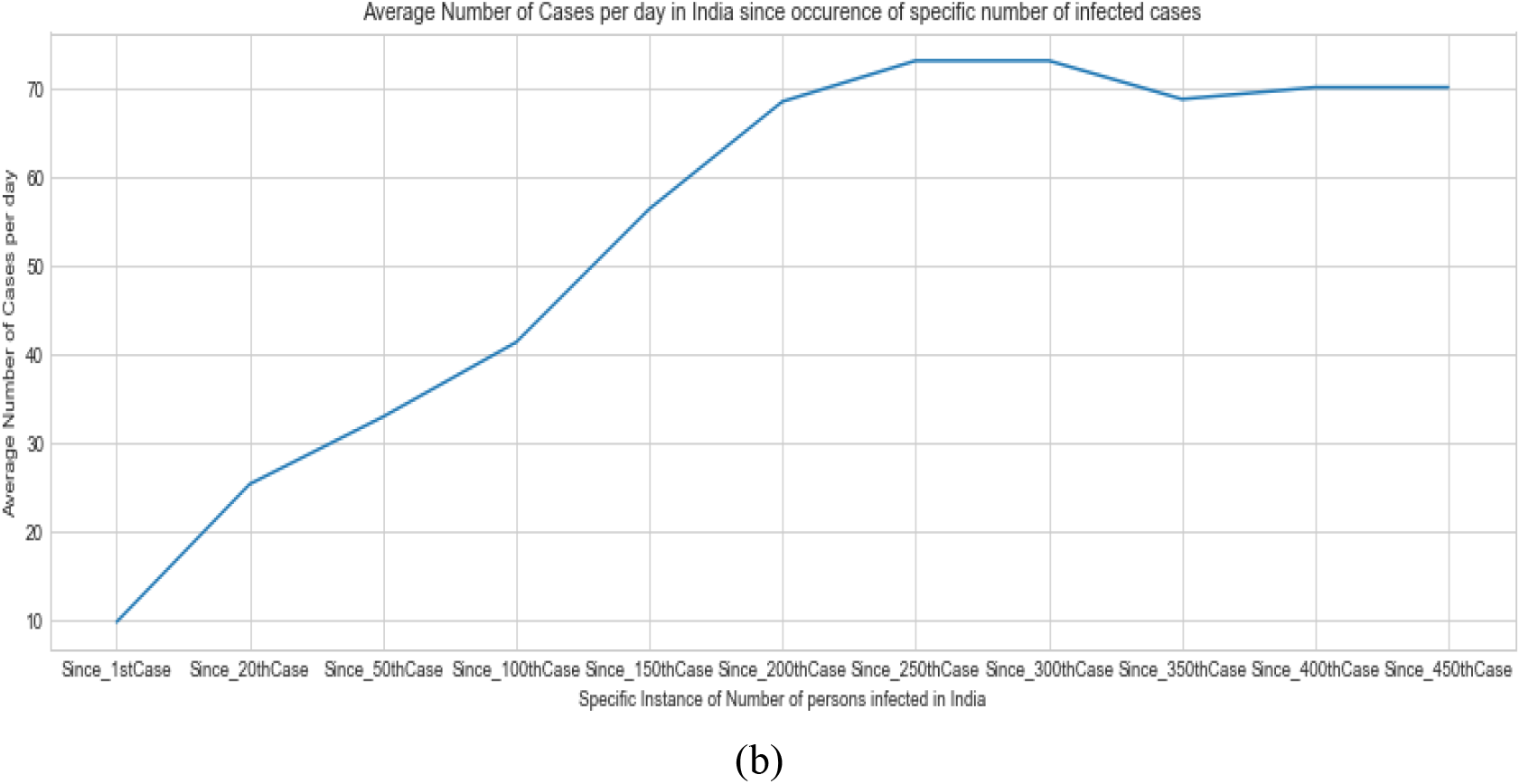
Trends of average number of cases reported due to COVID-19 in India. (a) Number of days to reach the specified range of infected people in India. (b) Average number of cases per day in India since occurrence of a specific number of infected cases

### B. Trends in Indian States

Indian states are witnessing different patterns with respect to the reporting of the COVID-19 patients. The trends are shown in Figure 3. As seen in the Indian states and union territories (UTs), the infected patients are getting recorded in more than 75% of the region of India. Almost 25 states and UTs reported at least one case of infected patient. Kerala and Maharashtra are at the top currently with more than 100 cases being reported by each state. Karnataka, Uttar Pradesh, Gujarat, Rajasthan and Delhi have reported patients in the range of 31-41, while Tamil Nadu, Haryana and Ladakh region have reported cases in the range of 11 to 30. Rest other regions have reported less than 10 cases. Out of these states, Maharashtra have reported 2 deaths while Karnataka, Gujarat, Delhi, Punjab, West Bengal, Bihar and Himachal Pradesh have reported 1 death each. Since the data is skewed at the moment, so the mortality rate of Bihar and Himachal Pradesh is highest ranging around 33%. However, currently the mortality rates are ranging from 1% to 3.5% respectively.

**Figure 3.**
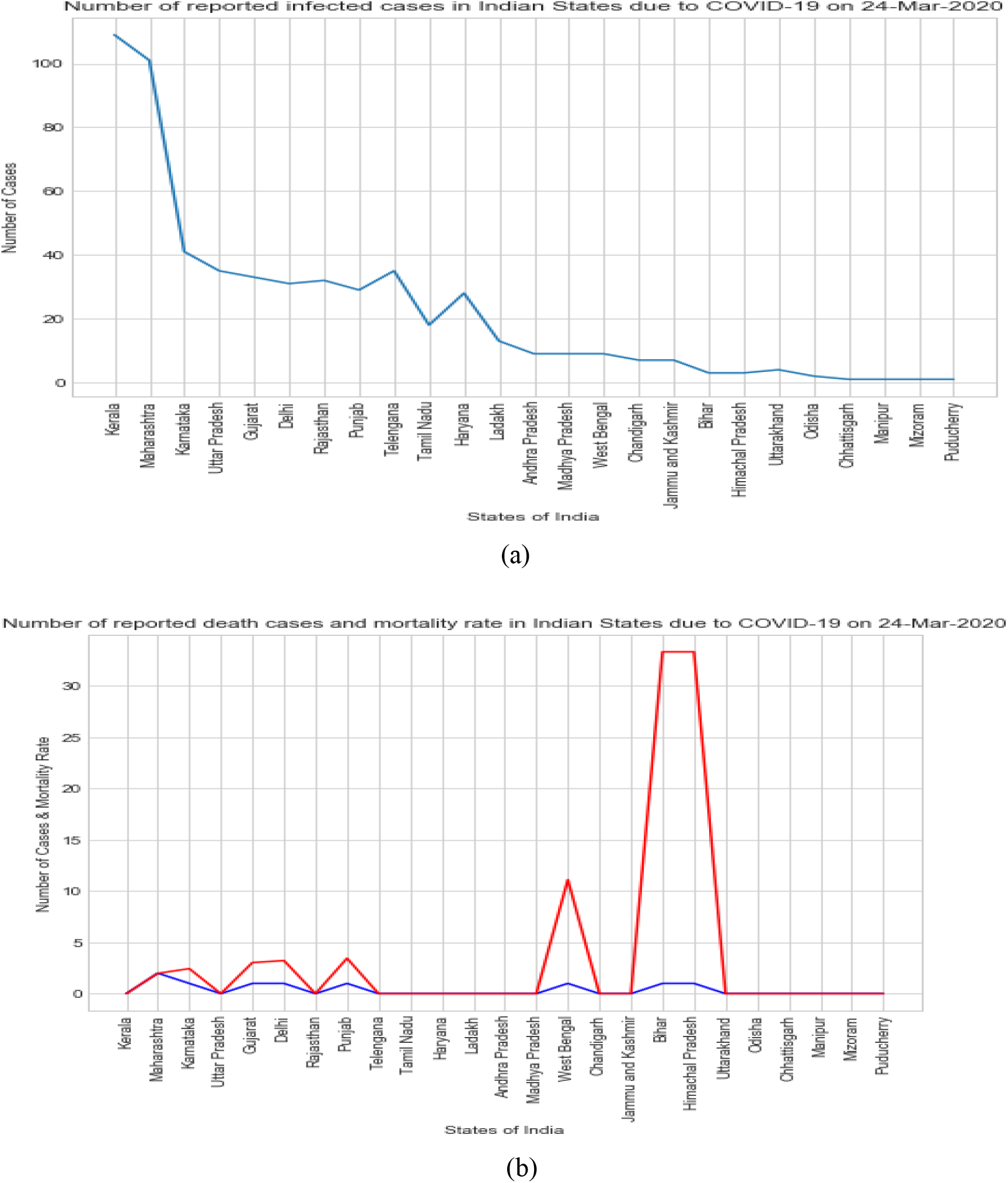
Trends of average number of cases reported due to COVID-19 in India. (a) Number of days to reach the specified range of infected people in India. (b) Average number of cases per day in India since occurrence of a specific number of infected cases

### C. Trends in South Asia

As per available data, India did not perform well in South Asian region on the COVID-19 outbreak till now. Comparing the Indian cases to the members of other SAARC nations, data indicates that India is better than Pakistan in terms of number of infected cases reported so far. India reported its first case much earlier than other nations in south Asian region, but due to massive population and high population density, the growth curve seems to be exponential right now.

As seen from Figure 4, Pakistan reported a steep rise in the number of COVID-19 cases in last 2 weeks reaching upto 900 patients as compared to 536 in India. The number of COVID-19 patients in Sri Lanka is close to 100, while those of Afghanistan and Bangladesh are less than 50. Bhutan and Nepal have less than 5 patients while Maldives has around 10 infected cases. The number of deaths are low in South Asia with India reporting around 9 deaths while Pakistan and Bangladesh reporting 7 & 4 deaths, respectively. Other nations are yet to report any deaths. India being the most populous country in South Asian region, the numbers of COVID-19 cases are rising. The Government of India has also taken an initiative to create a $10 million fund for combating COVID-19 outbreak in SAARC region [32]. With the help of neighboring countries, India is planning its combat strategies and has stopped all cross border activities for now.

**Figure 4.**
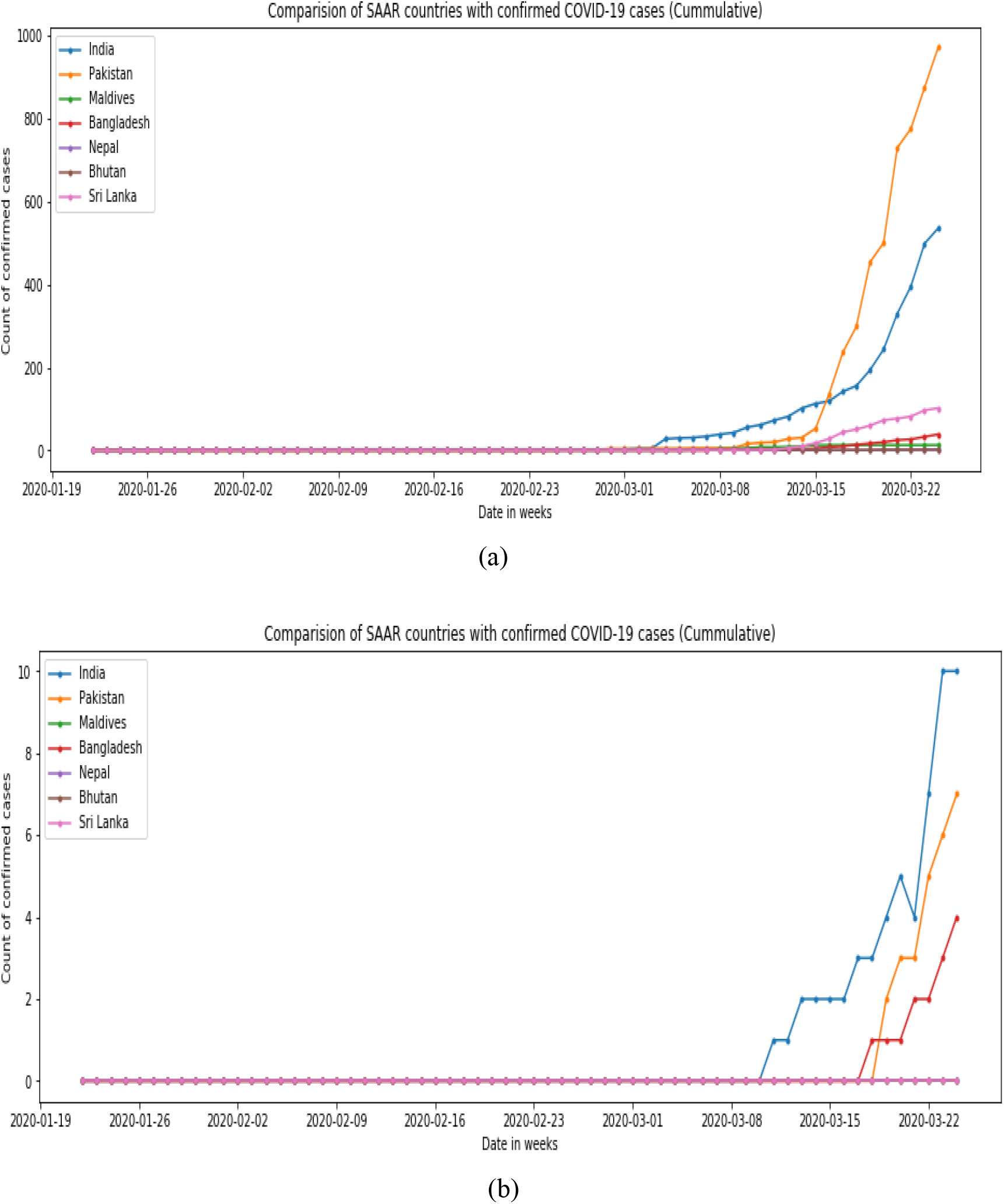
Trends of cases reported due to COVID-19 in SAARC countries. (a) Number of reported cases (cumulative). (b) Number of Deaths reported (cumulative)

### D. ARIMA modeling based Forecasts in India

India is in the growth stage but needs a lot of work towards maintaining the adequate resources and imposing restrictions on social gatherings to avoid contact base. With the number of infected people rising sharply in last one week, ARIMA model gives the predictive range of possible infectious patients in India. Figure 5 shows the natural log of the number of infected cases as there was an exponential upward trend seen in the original data.

**Figure 5.**
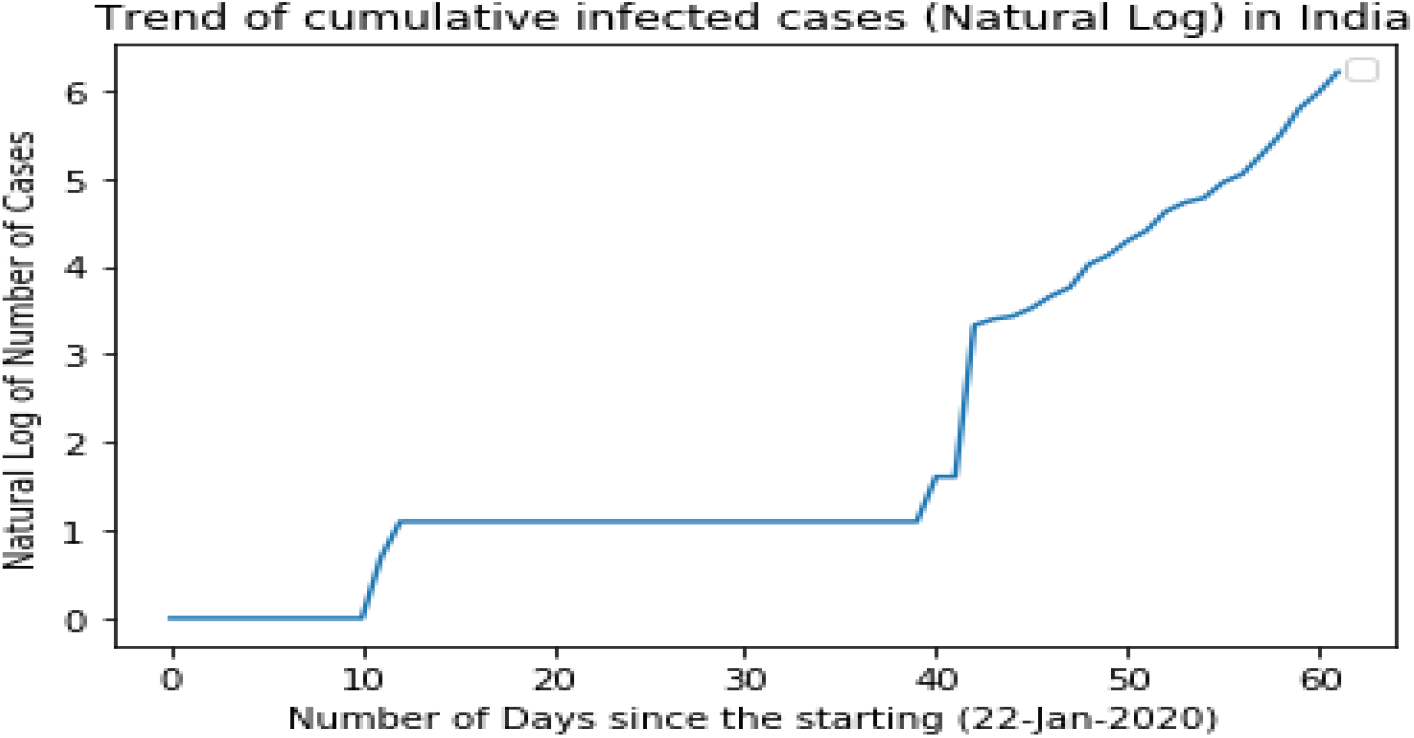
Trend of cumulative infected cases (Natural Log)

Even after performing the natural log transformation on the raw data, the upward trend was still visible. Since, this can hamper the stationarity of the series and can pose difficulties in fitting the ARIMA model, so exponentially weighted moving averages were applied to eliminate the trend and seasonality from the data. Figure 6 shows the rolling mean and standard deviations which makes the series stationary as confirmed by Dickey-Fuller test too.

**Figure 6.**
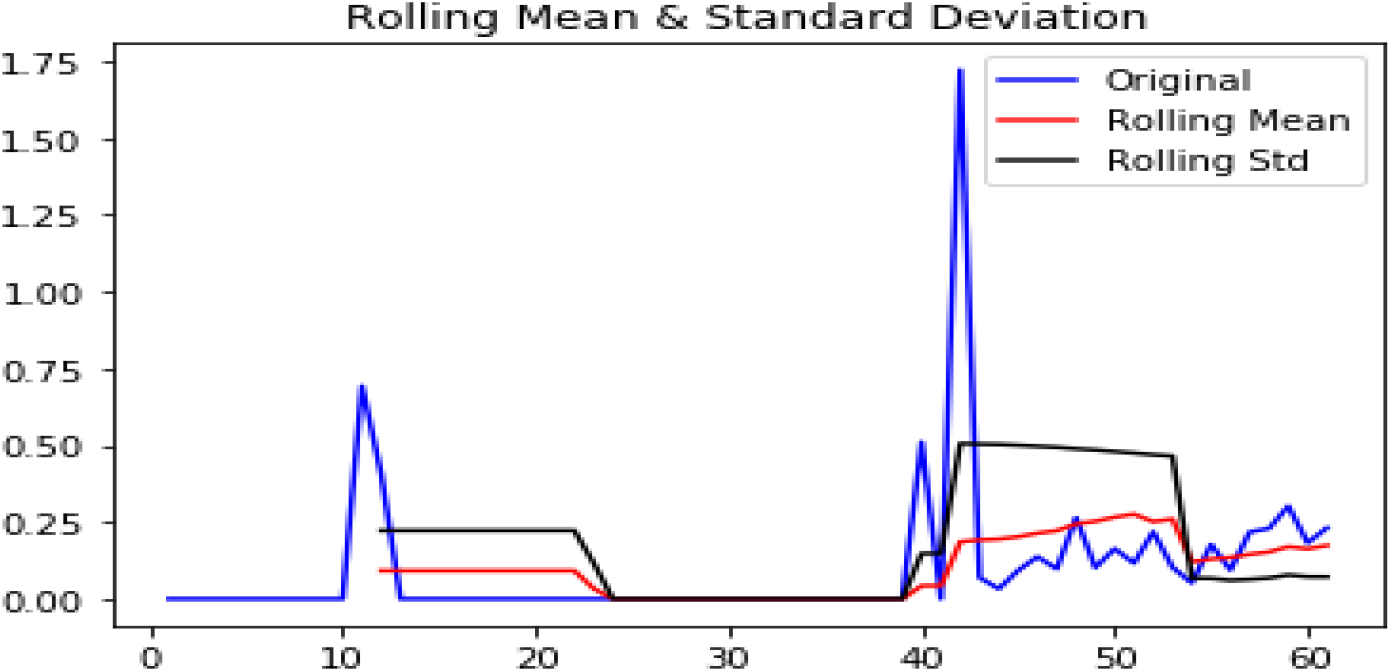
Series with rolling mean and standard deviation

As shown in Table 1, the first order auto-regressive component and second order moving average component were found to be statistically significant for the ARIMA(1,1,2) model. Therefore, model can be considered for the predictions.

**Table 1.** Model Summary for ARIMA(1,1,2) model

|  | Coef | Std Err | Z | P>\|z\| |
| --- | --- | --- | --- | --- |
| Const | 10.1403 | 7.123 | 1.424 | 0.161 |
| ar.L1.D | 0.6741 | 0.114 | 5.928 | 0.000 |
| ma.L1.D | -0.0247 | 0.105 | -0.236 | 0.816 |
| ma.L2.D | 0.7921 | 0.116 | 6.840 | 0.000 |

The graphs shown in Figure 7 depict the predictability of the model and spread over the original data. After removing the trends and seasonality from the log transformed data, the forecasted line follows the data line. Also, originally the log transformed data series has been well forecasted by the ARIMA model.

**Figure 7.**
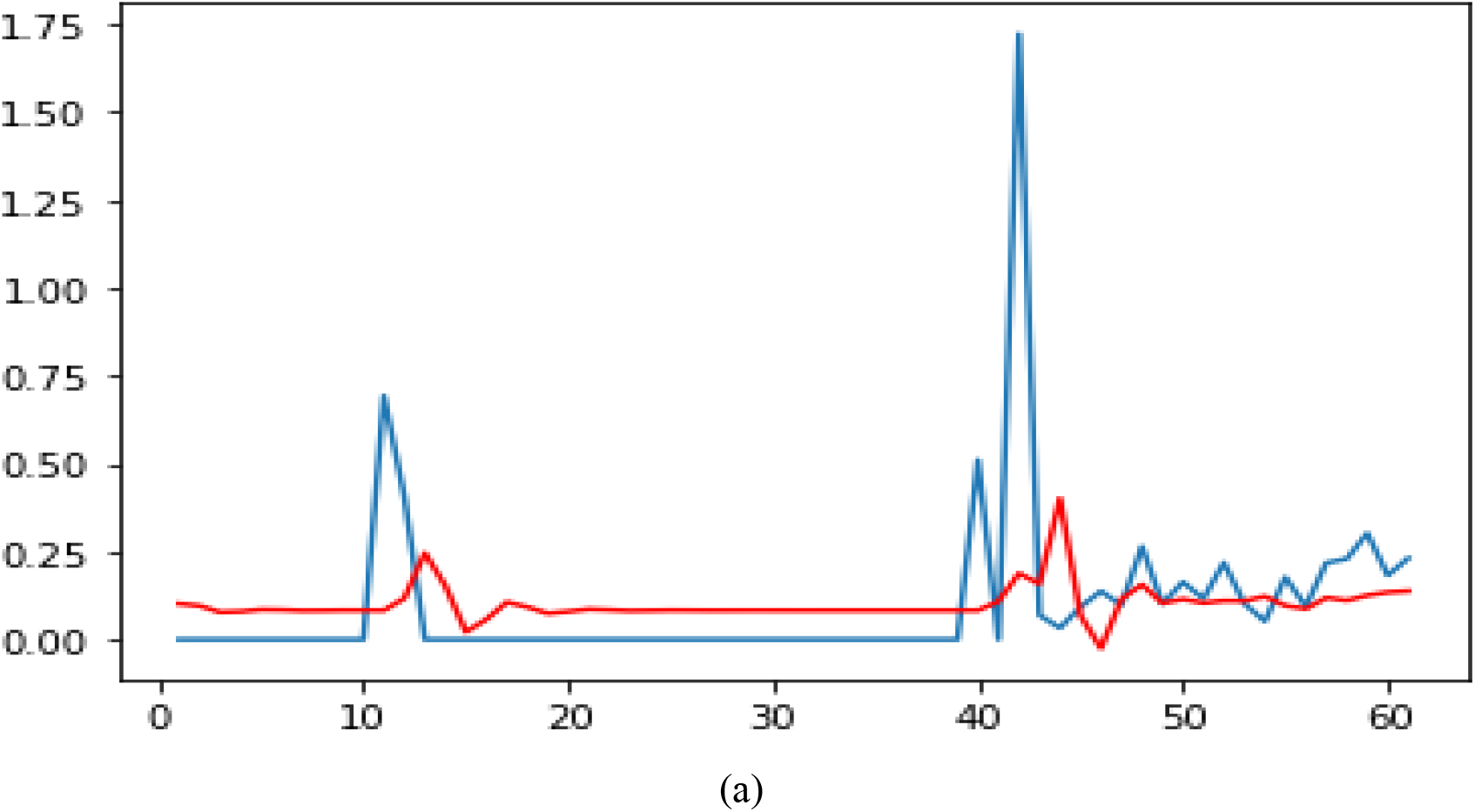
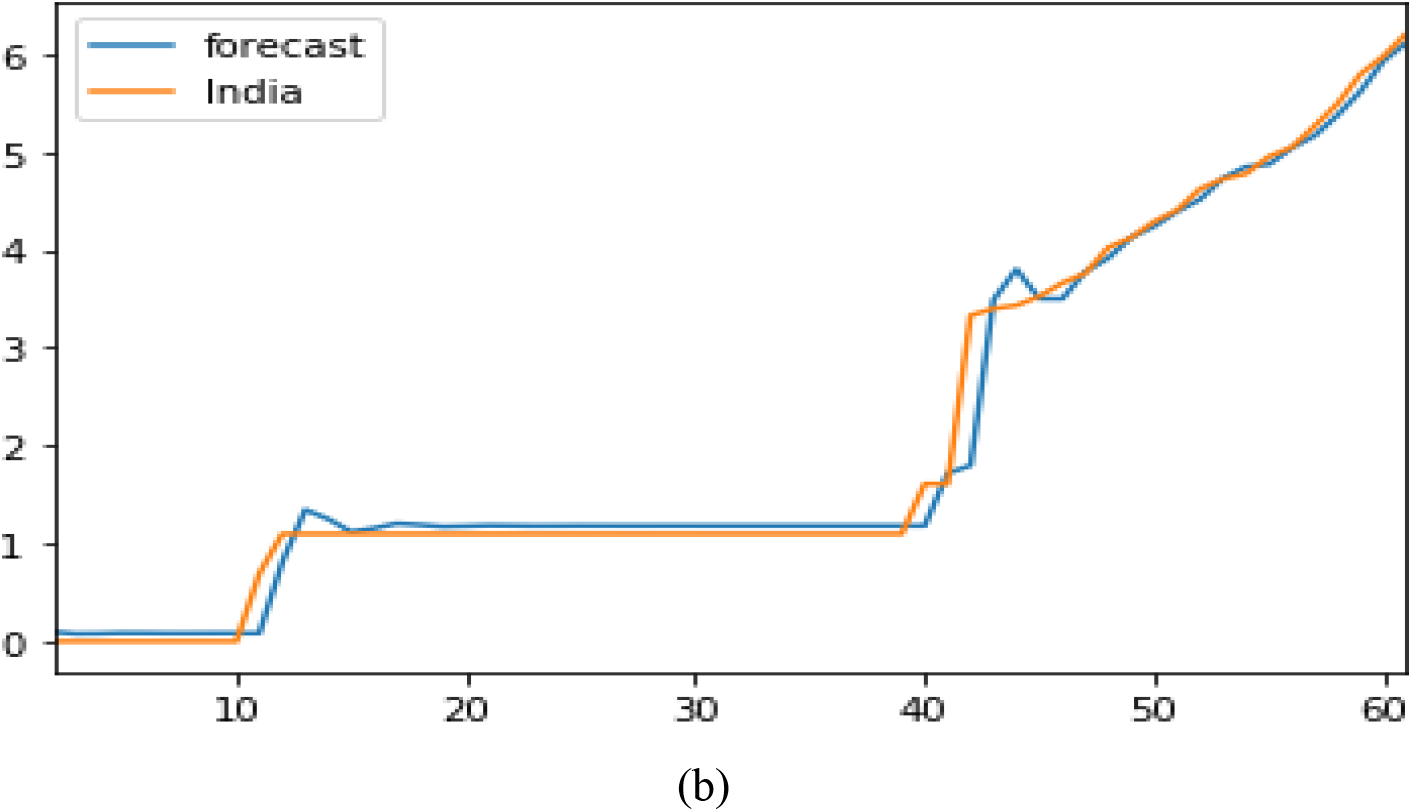
Forecast trends over the modelled data. (a) Shows the forecasted line in red and stationary data in blue. (b) Forecasted data line is shown in blue while the originally log transformed data is shown in Orange.

Figure 8 shows the future trend predictions which seem to be quite alarming at 95% confidence interval. Looking at the graph range for next 30 days, the optimistic scenario looks to be in control while the pessimistic scenario looks quite horrifying, with the average case being in the normal growth scenario.

**Figure 8.**
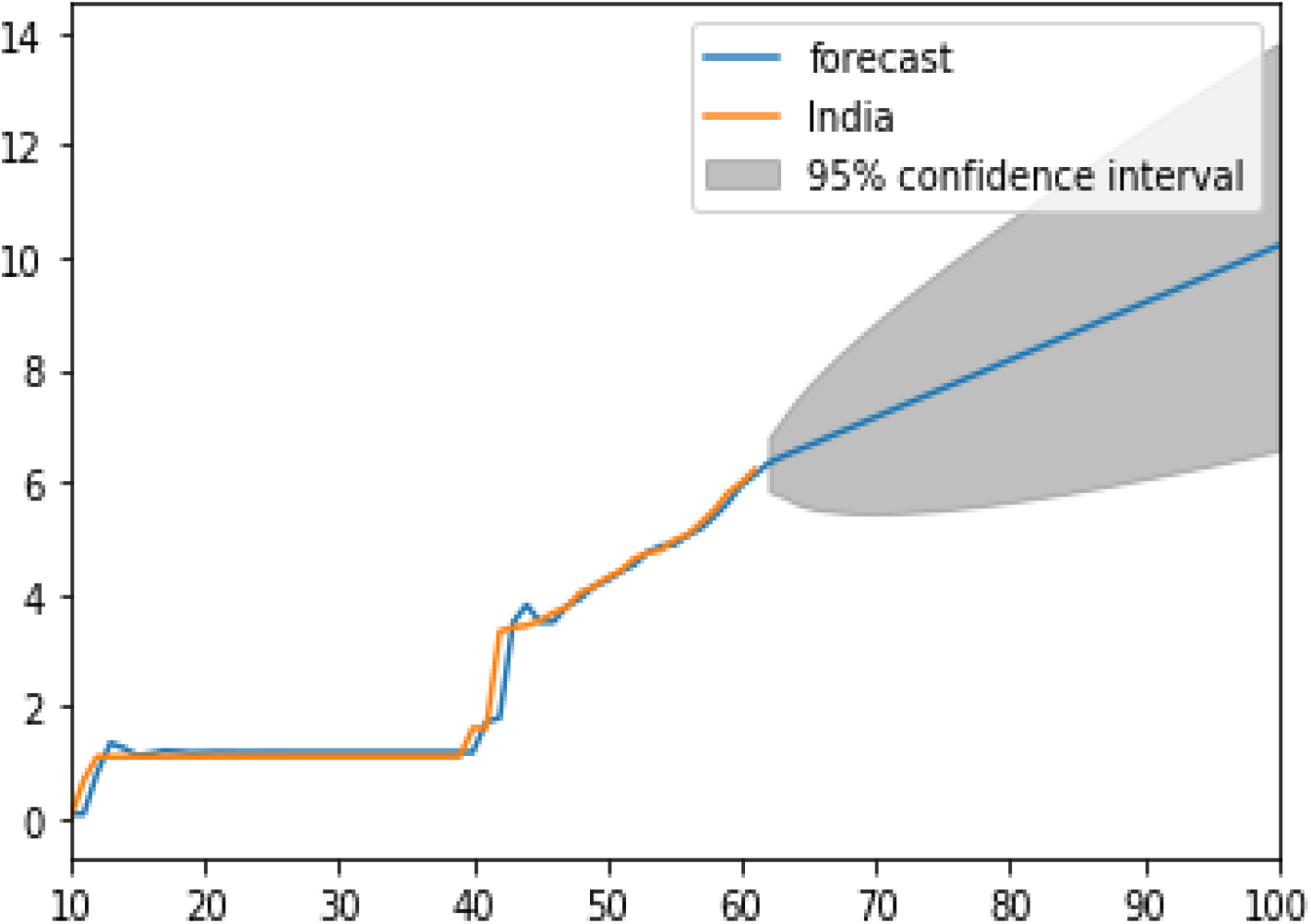
Graph showing the forecasted band at 95% confidence interval for the next 30-35 days.

The range for the predicted log values comes out to be around 13.8 in the worst case scenario while it remains around 6 in the best case scenario and hovers around 9.5-10 in the average case scenario. Transforming these numbers, it can be said that the expected number of cases may rise up to 700 thousand in the worst case scenario while it may remain controlled around 1000 patients only in next 30 days if strict measures are taken. On the average growth rate predictions, the cases may rise up to 7000 in next 30 days as compared to the current numbers of 536. Considering the mortality rate to be varying between 2-3% based on the early trends, the number of deaths is predicted to be in the range of 15,000-20,000 in pessimistic scenario. The accuracy of the model was found to be around 70% as the data points are less.

### E. Exponential Smoothing based Forecasts in India

Exponential Smoothing is one of the most widely used techniques for the forecasting of the trends on time series dataset. The results of the forecasting trends are shown in Figure 9.

**Figure 9.**
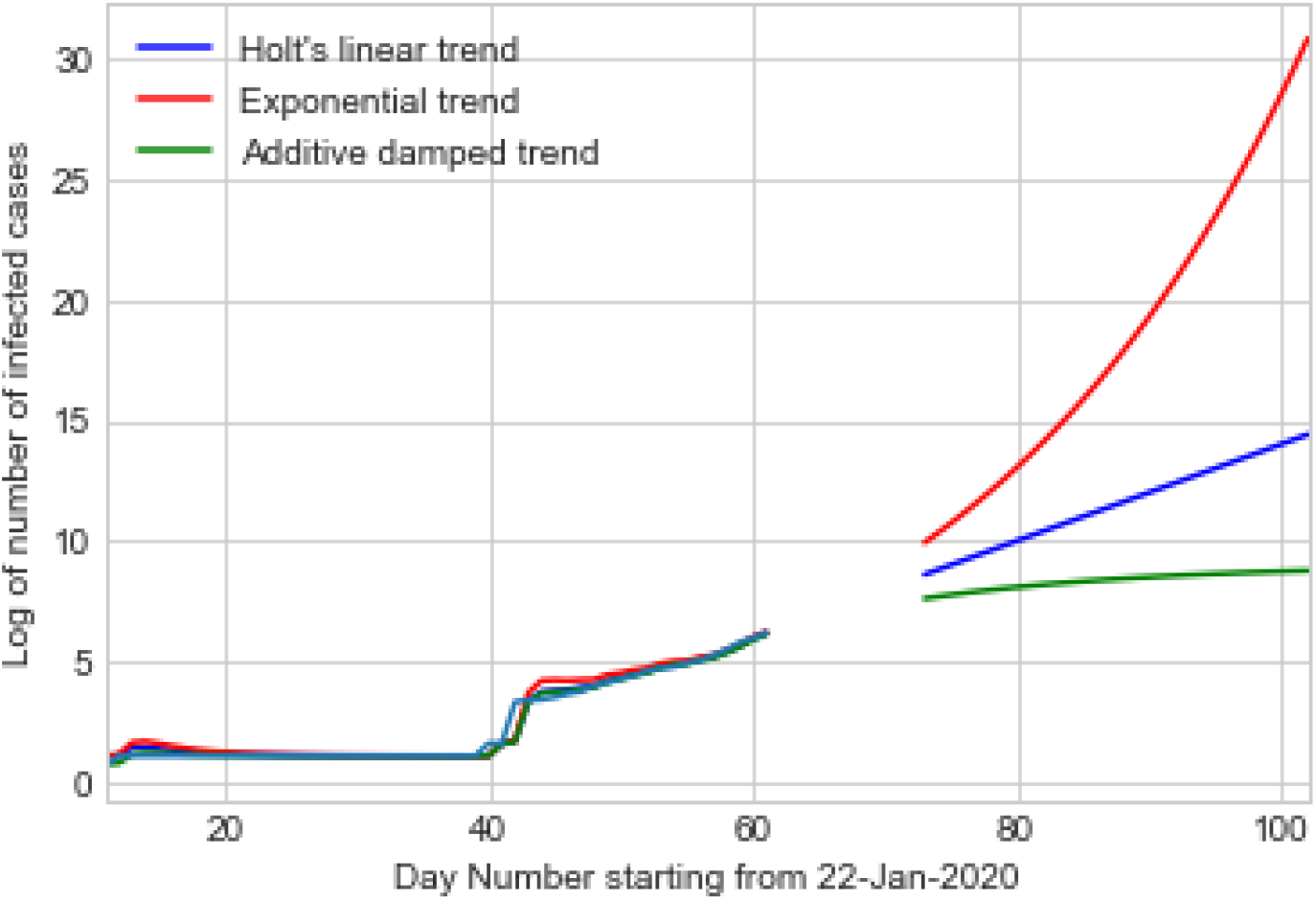
Graph showing the forecasted trends through Holt’s Linear trend, Exponential trend, and Additive damped trend for the 25-30 days in India.

The three methods used in the forecasting are all related to the Holt’s Linear Exponential Smoothing methods. As per the exponential trends, the number of infected persons may grow very quickly in next 30 days and my reach to more than 80 million infected cases without any intervention. Of course, the interventions are going on and a lot of people are recovering around the world and in India. However, the Holt’s Linear Trend is a realistic estimate of the situation in the current scenario and going by the forecasts, we may see around 3 million infected cases in India in the next 30 days or so. To control the wild predictions, Additive dampening method was also used to predict the forecast and this model depicted that with proper interventions and control, the figures may not reach beyond 8000-9000 infected cases.

### F. Infected Health Workforce

The risks of getting infections to the healthcare professionals around seriously ill people under the pressure of a pandemic are almost impossible to avoid. In hard-hit Italy, 20 percent of health-care professionals in the Italian region (more than 4800) have become infected with the virus, according to an update Friday in the Lancet medical journal [33]. In China, 3,387 health-care workers were infected by Feb. 24, almost all in Hubei province, the center of the outbreak, according to Chinese health authorities [33]. In Connecticut, at least 200 nurses have been sidelined from their duties and put in isolation due to lack of testing [34]. In Pittsfield, Massachusetts, 160 employees of Berkshire Medical Center have been quarantined due to exposure to the virus [35]. Older and vulnerable health care workers can help fight Covid-19 from a safer distance than they do today. Clinics across the U.S. have shifted to phone or virtual encounters; vulnerable health care workers could absorb some of this load to free younger ones with more robust immune systems to transition to clinical work in hospitals [35].

Within India, doctors and other health care professionals are yet to report any case of COVID-19 outbreak due to attending patients with the virus. But they are not averse to catching the infection from some other host. However, medical workers in India are facing logistic concerns with respect to their rented accommodations as most of the landlords of house rented to medical professionals are asking them to vacate the premise in order to avoid catching the novel coronavirus in their body. The governments of respective states are addressing the issue, but if healthcare workers do not report to the respective work locations, we may not be able to contain the drastic rise in the number of deaths and infected case due to COVID-19 outbreak. And looking at the global trends, it is important for the Indian workforce to exercise utmost precautions, guided by best medical practices and proper medical supervision.

### G. Infrastructure in India

The medical healthcare infrastructure is not one of the best in India. As per the report by The Print [36] media house, India has roughly 670 thousand hospital beds while 70 thousands ICU beds which are used for critical care. There are around 1.1 million doctors available for the treatment and a workforce of 2.05 million nurses available in the Indian region. The number of ventilators is estimated to be about 40,000 all over the country. The availability of beds per 1000 people is worse in India with just 0.5 beds available for every 1000 patients in Indian. This ratio is not evenly spread in different regions of India. The range of 4.5 beds per 1000 persons is estimated in different states of India with highest being around 4.7 and lowest being around 0.2 beds. Moreover, the testing kits are not sufficient enough to even cater 0.2 million of patients currently. These figures are surely discouraging given the projections of the number of patients due to COVID-19 outbreak in India.

The Government of India is coming up with quick financial packages for improving the infrastructure and procuring more number of testing kits and developing more number of testing labs. The tourism infrastructure and railway infrastructure has been planned for getting more number of quarantine zones where COVID-19 positive patients can be kept. Private hospitals and labs have come forward in support of the Government of India to overcome the challenge of weak healthcare infrastructure in India.

## IV. Discussions

The projected situation is very alarming. The results of the current study are in line with the predictions done by Indian Council of Medical Research [18] where 0.2-9 million people alone in Delhi have been projected to be infected in different scenarios with the help of hypothetical modeling implemented on a small dataset. Other authors have also projected cases in other countries like China and Italy using Time Series forecasting the accuracy have been found to be around 60-70% as is the case with the current study.

The projections in the current study are based on basic fundamentals of the Time Series Forecasting techniques but may become reality if necessary measures are not taken seriously. In this regard, the Government of India has already announced a complete lockdown of the country on 24^th^ March 2020 for 21 days to maintain social distancing among the citizens of the country. All the essential services including daily needs and medical store are open for limited accessibility and special phone lines have been setup to take any query related to symptoms and disease due to COVID-19. This should have a huge impact on the flattening the curve rather than making it grow exponentially. Since the projected R_o_ value has been predicted between 1.5 and 4 by the ICMR [18], it is imperative that people should stay within their homes. In fact strict lockdown in countries like China and Singapore is yielding results and curves are flattening with respect to the new cases adding to the tally.

Prior to this, the Government of India also announced lockdown of domestic travel system including interstate road transportation, rail travels and flight travels. Most of the inbound and outbound international flights have been cancelled due to the prevention of the outbreak. And all the passengers coming to India from abroad are being kept under mandatory 14 day quarantine.

The entire world, including India, was taken by surprise and countries did not get any time for elaborate preparation. Social and physical distancing is a good move by the government but basic needs of different sections of the society should also be addressed. Welfare of stranded, poor, daily workers should also be addressed by government, corporates, and NGOs with government funding. The finance ministry has recently announced relief packages for the marginalized communities, but implementation at ground level needs careful and risk free planning amidst lockdown period. Government should also enhance testing facilities, hospital support, and personal hygiene on a war footing as exponential growth may not give a lot of time to react for the various officials and departments. Specific R&D centers in the country should work to quickly create the best possible solutions for cure like vaccines, oral medicines, Ayurveda substitutes, preventive kits, apart from focusing on producing hand sanitizers & masks. A lot of companies are in race to test the vaccine but yet the situation looks troublesome for now.

In fact at the global level, WHO should immediately initiate joint R&D for cure by pooling necessary resources from different continents and regions. Necessary funds should be allocated in combating the disease and region specific advisory should be announced on case to case basis.

## V. Conclusion

To conclude, the current study focused on presenting trends and forecast in the Indian region with respect to the outbreak of COVID-19. Although the growth rate is not at par with growth rate at World Level, but the situation looks dangerous as India is heading towards exponential growth. Through two different time series forecasting models, the projected patients are reaching in millions in next 30 days. With relatively weak healthcare infrastructure, it would be difficult to contain the outbreak of this disease without drastic steps by the Government. In addition to more stringent lockdown mechanisms for physical distancing, patient testing, isolation & medical treatment mechanisms should be taken up on a war footing for controlling this pandemic in India.

The projections in this study are still in early phases as the historical data is very less for the model to be really accurate. It is only in the last 10 days, that the cases have risen in India. So the training of the model may not be really accurate for now. But the prediction model will certainly improve with more of medical and demographic data in the existing models. However, even if the projections are 60-70% accurate, then also it would be really difficult days for the country. In future, the study on pre-lockdown and post-lockdown can be conducted to see if social distancing in India is actually working.

## Data Availability

Data is publicly available on the web repository of John Hopkins University.

https://github.com/CSSEGISandData/COVID-19

## Acknowledgments

The authors would like to thank the John Hopkins University for publicly releasing the updated datasets on the number of infected cases and death cases occurring worldwide due to COVID-19.

## About Authors

**Dr. Rajan Gupta** is a Research and Analytics professional and is currently associated with University of Delhi in India as Assistant Professor. He has done his PhD from University of Delhi in the area of Application of Data Science in Improving E-Governance in Developing Nations. He has written two books on E-Governance in India and has published various papers at national and international forums. He is an active consultant to a lot of organizations in the area of Bio-statistics and Data Science.

**Dr. Saibal K Pal** is from DRDO, a R&D organization under the Ministry of Defence, India. He has done his PhD from University of Delhi and has been involved in lot of projects related to implementation of E-Governance in India. His area of research includes security, information systems and E-Governance. He has published two books on E-Governance in India and has published various papers at national and international forums.

## Notes

### Competing Interest Statement

The authors have declared no competing interest.

### Funding Statement

Not Applicable

